# IgG4 Neutralization and Sustained Total IgG Fc-Effector Functions Following Repeated SARS-CoV-2 mRNA Vaccination

**DOI:** 10.1101/2025.09.30.25336755

**Authors:** Paulina Kaplonek, Harry Bertera, Diana Lee, Deniz Cizmeci, Wen-Han Yu, Kai Wu, Spyros Chalkias, Rahnuma Wahid, Darin Edwards, Galit Alter, Carole Henry

**Author notes:** Affiliation at the time of the study (Galit Alter).

## Abstract

Detailed characterization of the antibody profile induced by SARS-CoV-2 mRNA vaccines has shown that repeat dosing boosts all IgG subclasses, with a notable emergence of antigen-specific IgG4 antibodies. While the IgG4 subclass is traditionally associated with limited Fc-effector functions, its role in SARS-CoV-2 mRNA vaccine-induced immunity remains unclear. This study tracked IgG subclass dynamics, IgG Fc-mediated functions and neutralization following immunization with two or three doses of mRNA-1273. We observed robust Spike-specific IgG1 and IgG3 responses after the primary series (two doses) and booster dose, with a significant increase in IgG4 responses after repeated dosing. Despite this rise in spike-specific IgG4 antibodies, strong Fc-effector functions were maintained at the overall IgG level, including ADCP, ADNP, and ADCD, with no antagonism from IgG4 antibodies. Additionally, IgG4 antibodies exhibited enhanced affinity and potent neutralization, complementing IgG1-driven responses. These findings suggest that the IgG4 response induced by mRNA-1273 vaccination co-evolves with other subclasses to enhance Fab- and Fc-mediated immunity, underscoring the ability of mRNA vaccines to induce a balanced, multi-subclass immune response optimized for durable protection and robust functionality.

## Introduction

The COVID-19 pandemic prompted unprecedented progress in vaccine development, leading to the emergence of novel vaccine platforms, including mRNA vaccines, which were among the first to receive emergency use authorization due to their robust efficacy and safety profiles demonstrated in Phase 3 trials (Pfizer 2020; Moderna 2020). Despite the emergence of SARS-CoV-2 variants capable of evading neutralizing antibodies, vaccination continues to provide significant protection against severe outcomes, including hospitalization and death (Chen X, 2021; Mistry P, 2022). Booster or updated vaccination can further enhance protection against disease (Lin DY, 2023).

Antibodies serve as a key correlate of protection for most approved vaccines (Plotkin SA, 2010; Plotkin SA, 2023). Following vaccination, early IgM responses are typically induced as part of the primary humoral immune response. These IgM antibodies subsequently undergo class-switch recombination, predominantly to IgG1 or IgG3 for viral vaccines in the context of systemic immunity or to IgA in mucosal tissues, depending on the vaccine platform and the immunological environment (Pollard A.J., 2021). Unlike most vaccines that elicit IgG1-dominated responses, SARS-CoV-2 mRNA vaccines have been associated with a notable increase in IgG4 antibodies after repeated doses. These antigen specific IgG4 responses exhibited higher avidity, but at the same time were associated with reduction of some Fc-effector functions (Irrang P, 2023; Kalkeri R, 2024; Kiszel P, 2023). Because of these properties, IgG4 antibodies are generally considered to be anti-inflammatory and often associated with reduced immune responses. Similar IgG4 responses have been observed with natural infection (Davis CW, 2019) as well as other vaccines, such as pertussis vaccines (Hendrikx LH, et al.). The induction of IgG4 antibodies after repeated SARS-CoV-2 mRNA vaccinations is a complex phenomenon with potential implications for vaccine strategy and immune response. Ongoing studies are needed to clarify the significance of this shift and its impact on both protective immunity and immune regulation.

In this study, we tracked the evolution of SARS-CoV-2-specific antibody responses and their functional properties following primary mRNA-1273 immunization and booster series in healthy adults. We observed that the rise of IgG4 antibodies co-evolved with IgG1 and IgG3 responses, which exhibited robust Fc-mediated effector functions. IgG4 antibodies demonstrated high affinity, potent neutralization capacity, and complemented the IgG1-dominated response by contributing to antigen binding while potentially modulating inflammatory Fc-effector functions. Previous studies have suggested that IgG4 responses after repeated mRNA vaccination are associated with reduced Fc-effector activity and may direct immunity toward an anti-inflammatory profile (Irrang P, Kalkeri R, Kiszel P). In contrast, our findings indicate that SARS-CoV-2–specific IgG response, in the context of mRNA-1273 immunization, retains robust Fc-mediated functional activity. This suggests that the IgG4 antibody response evolves in tandem with IgG1-driven immunity, providing additional functionality that may enhance protection against COVID-19 following mRNA-1273 vaccination (Figueroa AL et al., 2025; Kopel H et al., 2024; Baden L et al., 2024; Berthaud V et al., 2024; Figueroa A et al., 2024; El Sahly H et al., 2021; Gilbert P et al., 2022).

## Methods

### Cohort

This is a Phase 3 study, Part C (Booster Dose Phase) of randomized, stratified, observer-blind, placebo-controlled study (NCT04470427) to primarily evaluate the efficacy, safety, and immunogenicity of mRNA-1273 SARS-CoV-2 vaccine in adults aged ≥18 years (Follmann D. 2024; Zhang, B., 2024; Chalkias S, 2022; Sahly HM. 2022). Participants who completed two-doses primary series of IM injection of 100 μg mRNA-1273 at least six months prior, received a single booster dose of 50 μg mRNA-1273. The mRNA-1273 is a lipid nanoparticle containing mRNA encoding the spike glycoprotein of the original strain of SARS-CoV-2.

### Luminex

Serum samples were run in a customized Luminex assay to quantify the relative concentration of antigen-specific antibody isotype and subclass profiles. Carboxylated magplex-microspheres (Luminex) were coupled to antigens SARS-CoV-2 WT Spike protein using covalent NHS-ester linkages by EDC and NHS (Thermo Fisher Scientific) as described previously (Brown EP, et al., 2017). To form immune complexes, appropriately diluted serum (1:100 for IgG2/3, 1:500 for IgG1, and 1:1000 for all other FcγRs), was added to the antigen-coupled microspheres, and plates were incubated overnight at 4ºC, shaking at 700 rpm. The following day, plates were washed with PSA containing 0.1% BSA 0.02% Tween-20. PE-coupled mouse anti-human detection antibodies (Southern Biotech) were used to detect antigen-specific antibody binding. For the detection of FcγR binding, Avi-Tagged FcγRs (Duke Human Vaccine Institute) were biotinylated using BirA500 kit per manufacturer’s instructions. Biotinylated FcRs were tagged with PE and added to immune complexes. Fluorescence was acquired using an Intellicyt iQue, and relative antigen-specific antibody titer and FcγR binding is reported as Median Fluorescence Intensity (MFI).

### Antibody-dependent cellular phagocytosis (ADCP)

THP-1 cells (American Type Culture Collection, ATCC) were used in phagocytic assays were grown in RPMI-1640 (Sigma Aldrich) media supplemented with 10% fetal bovine serum (FBS) (Sigma Aldrich), 5% penicillin/streptomycin (Corning, 50 μg/mL), 5% L-glutamine (Corning, 4mM), 5% HEPES buffer (pH 7.2) (Corning, 50 mM) and 0.5% 2-Mercaptoethanol (Gibco, 275 μM). Cells were maintained at a concentration of 2.5×10^5^ cells/ml. Antibody-dependent cellular phagocytosis was measured using a flow cytometry-based phagocytic assay (Ackerman ME, et al., 2011). Briefly,1.0 μm, yellow-green fluorescent (505/515) FluoSpheres NeutrAvidin (Thermo Fisher Scientific) were coated with biotinylated SARS-CoV-2 WT Spike protein, incubated with serum samples diluted 1:100, and breastmilk diluted at 1:10. The ability of samples to drive uptake of antigen-coated beads by THP-1 cells after overnight incubation was assessed by flow cytometry using the iQue (Intellicyt). Phagocytic scores were calculated as follows: (% yellow green+ cells x yellow-green MFI)/100. A pool of serum samples collected from SARS-CoV-2 infected patients was used as a positive control, and a pool of serum samples from non-infected patients or 1x phosphate-buffered saline (PBS) alone was used as a negative control. Samples were run in duplicate.

### Antibody-dependent neutrophil phagocytosis (ADNP)

Antibody-dependent neutrophil phagocytosis was measured by a flow cytometry-based assay (Karsten CB, et al., 2019). Briefly, serum samples were diluted at 1:100, and breastmilk was diluted at 1:10. Samples were then allowed to form immune complexes with 1.0 μm yellow-green fluorescent (505/515 nm) FluoSpheres NeutrAvidin (Thermo Fisher Scientific) beads coated with biotinylated SARS-CoV-2 WT Spike protein. White blood cells were isolated from whole blood using ACK Lysing Buffer (Thermo Fisher Scientific) to lyse red blood cells at room temperature (1:10). After lysis, remaining cells were counted and resuspended at 2.5×10^5^ cells/ml in RPMI-1640 (Sigma Aldrich) media supplemented with 10% fetal bovine serum (FBS) (Sigma Aldrich), 5% penicillin/streptomycin (Corning, 50 μg/mL), 5% L-glutamine (Corning, 4 mM), and 5% HEPES buffer (pH 7.2) (Corning, 50 mM). Cells were then incubated with bead and antibody mixture for 1 hour at 37ºC and then stained with Pacific Blue-conjugated anti-CD66b (BioLegend, clone: UCH71, 2 μg/mL) in PBS for 15 minutes at room temperature in the dark. Finally, cells were fixed in 4% paraformaldehyde (PFA) and phagocytosis of beads by CD66b^+^ cells was measured by flow cytometry using the iQue (Intellicyt). A pool of serum samples collected from SARS-CoV-2 infected patients was used as a positive control, and a pool of serum samples from non-infected patients or 1xPBS alone was used as a negative control. Samples were run in duplicate. Data reported is an average of data points collected from two donors.

### Antibody dependent complement deposition (ADCD)

Antibody-dependent complement deposition (ADCD) was performed as previously described (59). In brief, SARS-CoV-2 antigens were coupled to magnetic Luminex beads (Luminex Corp) using carbodiimide-NHS ester chemistry (Thermo Fisher Scientific). The antigen-coated beads were incubated with serum samples for 2 hours at 37°C to form immune complexes, followed by washing to remove unbound immunoglobulins. Guinea pig complement (Cedarlane), reconstituted in gelatin veronal buffer with calcium and magnesium (GBV++, Boston BioProducts), was then added to the immune complexes to initiate complement activation. Complement component 3 (C3) deposition was detected using a fluorescein-conjugated anti-C3 goat IgG detection antibody (Mpbio). Complement deposition was quantified by measuring mean fluorescent intensity (MFI) via flow cytometry.

### Avidity/Affinity assay

The IgG affinity index was assessed by the Urea Luminex assay. General Luminex assay protocol described above was used with additional treatment step. the samples were treated for 10 minutes at room temperature with either 2M NH*SCN (Sigma-Aldrich) or phosphate-buffered saline (PBS) containing 1% BSA. After additional washing steps, the samples were incubated with PE-conjugated goat anti-human IgG. The mean fluorescence intensity (MFI) values were subsequently acquired using an FM3D instrument. The affinity index (AI) was calculated using the formula: *affinity index (%) = ((MFI NH4SCN) /(MFI PBS))* 100%*

### Neutralization

Serum samples were analyzed for nAb responses via vesicular stomatitis virus (VSV)-based pseudovirus neutralization assay (PsVNA). Codon-optimized full-length WT (D614G) spike genes were cloned into a pCAGGS vector. The recombinant VSVΔG-based SARS-CoV-2 pseudovirus was generated by transfecting BHK-21/WI-2 cells with the spike expression plasmid and subsequently infected by VSVΔG-firefly-luciferase as described previously [26]. For neutralization assay, A549-hACE2-TMPRSS2 cells were used as target cells. Serum samples were heat-inactivated for 45 minutes at 56°C, and serial dilutions were made in Dulbecco’s Modified Eagle Medium supplemented with 10% fetal bovine serum. The diluted serum samples or culture medium (serving as virus-only control) were mixed with VSVΔG-based SARS-CoV-2 pseudovirus and incubated at 37°C for 45 minutes. The inoculum virus or virus-serum mix was subsequently used to infect A549-hACE2-TMPRSS2 cells for 18 hours at 37°C. At 18 hours after infection, equal volume of One-Glo reagent (Promega) was added to the culture medium for readout; the percentage of neutralization was calculated based on relative luminescence units (RLU) of the virus-only control and was analyzed using four-parameter logistic curve (Prism v.9).

### Analysis

Data was visualized using RStudio (R ver. 4.4.1) with statistical significance calculated using Wilcoxon ranked test (rstatix ver. 0.7.2). Luminex binding assay, Antibody-dependent complement deposition, and Affinity binding assay data was Log10 transformed for visualization purposes. Tertile ranking was determined using ntile function (dplyr ver. 1.1.4) to determine 3 buckets of ranked values. Phagocytosis score is an arbitrary unit that is calculated in iQue3 software (iQue ForeCyt ver. 9.1) using the following formula, *(GeoMean BL4-H bead positive cells * % bead positive cells) /100,000*. Data points generated from the neutralization assay were normalized to the concentration of the purified IgG (1 ug/mL) and Log_2_ transformed for visualization.

## Results

### mRNA-1273 Immunization Induces Dynamic SARS-Cov-2-Specific IgG Subclass Response

To better understand serum antibody isotype and subclass dynamics after repeated doses of mRNA-1273 vaccination, we profiled the mRNA-1273-induced humoral response in SARS-CoV-2 naïve individuals at baseline, after the primary vaccination series (post-prime), prior to a booster dose (pre-boost), and following a third vaccine dose (post-boost) (**Figure 1A**). Robust IgG1 responses targeting the wild-type (WT) SARS-CoV-2 Spike antigen included in the vaccine were observed following the primary series. These responses partially contracted over the next 11–14 months but were boosted to levels comparable to those observed after the primary series following the third vaccine dose (**Figure 1B**). Spike-specific IgA responses were induced after the primary series, declined over time, and were boosted again following the third dose (**Figure 1C**). In contrast, Spike-specific IgM responses were induced at the lowest levels among all isotypes after the primary series (log_2_ fold change IgG1 = 8.37, IgA = 4.53, and IgM = 1.92), declined to near-baseline levels over time, and showed only a slight increase after the booster (**Figure 1D**).

**Figure 1.**
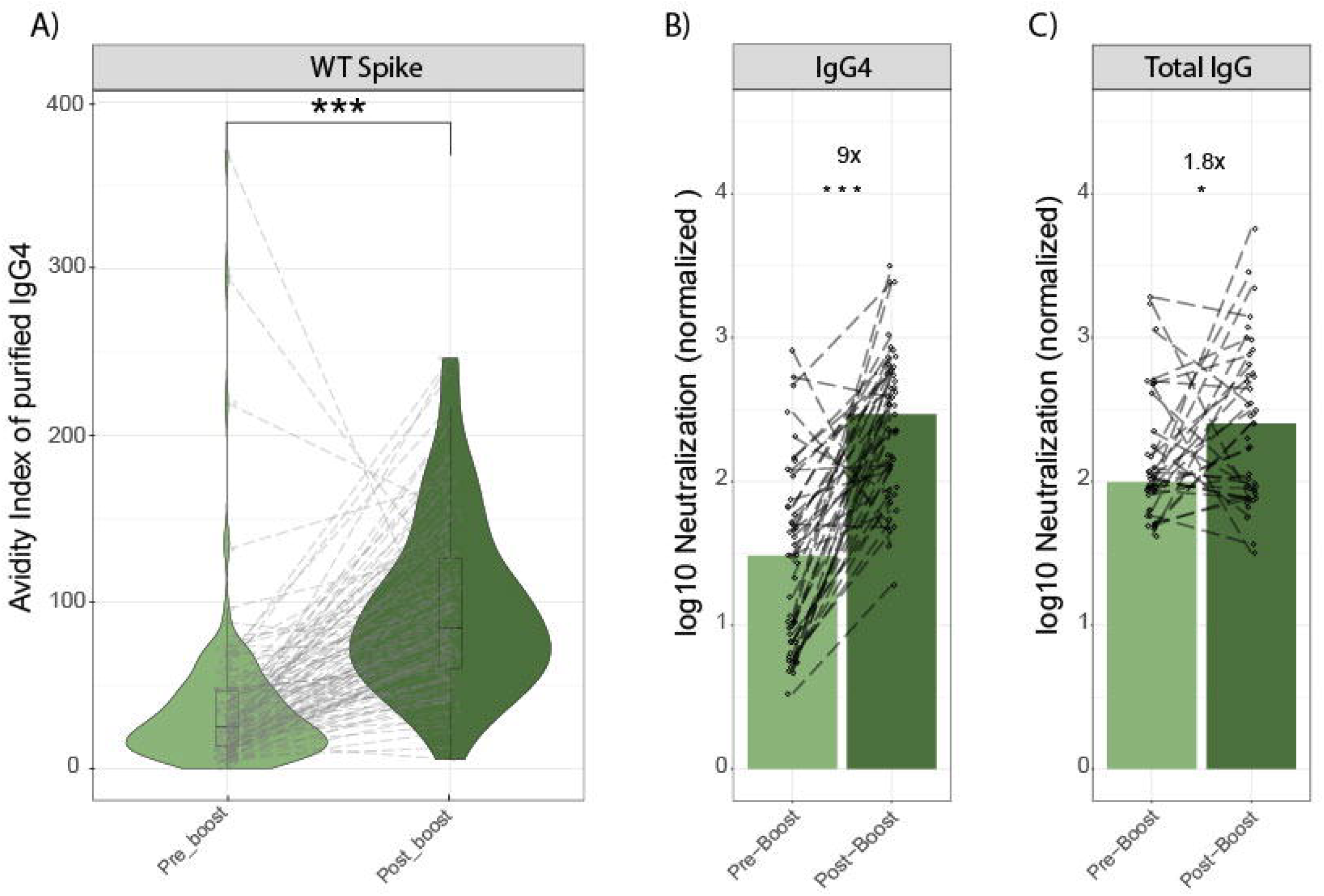
Evolution of Spike-specific antibody responses and subclass selection after repeated doses of the mRNA-1273 SARS-CoV-2 vaccine. **(A)** Immunization schedule: Healthy individuals (n = 79) were vaccinated with two doses of 100 µg mRNA-1273 (primary series, 29 days apart) and boosted 11–14 months later with 50 µg of mRNA-1273. Blood samples were collected at baseline, post-prime (29 days after the second dose), pre-boost (11–14 months after the primary series), and post-boost (29 days after the booster).Violin plots depict the levels of WT SARS-CoV-2 Spike-specific IgG1 **(B)**, IgA **(C)**, and IgM **(D)**, as well as IgG subclasses IgG2, IgG3, and IgG4 **(E)** at the specified timepoints. Dotted lines connect individual participants across all timepoints. The IgG1 to IgG4 antibody ratio **(F)** and IgG3 to IgG4 antibody ratio **(G)** analyses illustrate the dynamic shifts in antibody subclass distributions. The dotted line indicates an equivalent ratio between the two subclasses. Mann-Whitney U-tests, corrected for multiple comparisons using the Benjamini-Hochberg (BH) method, were applied. Adjusted p-values are denoted as follows: p < 0.001 ***, p < 0.01 **, p < 0.05 *.

To further characterize the humoral response, we analyzed changes in other IgG subclasses (IgG2, IgG3, and IgG4) (**Figure 1E**). Spike-specific IgG2 responses increased slightly after the primary series, significantly declined over time, and were robustly boosted after the third dose (log_2_ fold change baseline to post-prime = 3.2, and pre-boost to post-boost = 4.44). Spike-specific IgG3 responses were strongly induced after the primary series, but they declined to baseline levels and were not boosted following the third immunization (log_2_ fold change baseline to post-prime = 5.21, and pre-boost to post-boost decrease = 2.27). Interestingly, spike-specific IgG4 responses were not observed immediately after the primary series but continued increasing over time, with the highest levels being observed after the third dose (log_2_ fold change baseline to post-prime = 1.08, post-prime to pre-boost = 3.13 and pre-boost to post-boost = 4.43). To evaluate the balance and dynamics of IgG subclasses, we performed a ratio analysis to compare their relative abundances across different time points. The ratio of IgG1 to IgG4 revealed a dominance of the spike-specific IgG1 response over spike-specific IgG4 after the primary series, with a more balanced subclass distribution observed pre- and post-boost (**Figure 1F**). The IgG3-to-IgG4 ratio showed that although spike-specific IgG3 responses initially dominated over spike-specific IgG4 post-prime, IgG4 levels surpassed IgG3 over time and following the booster dose (**Figure 1G**). This analysis describes the dynamic shifts in antibody subclass distributions, highlighting distinct post-prime and post-boost antibody binding profiles, marked by a robust IgG1/IgG3/IgA response after the primary series that is replaced with a strong IgG1/IgG2/IgG4/IgA response after the boost.

### mRNA-1273 Vaccine Triggers Immune Response with Robust Fc-Mediated Antibody Effector Functions

Several studies have suggested that IgG4 has a reduced capacity to engage Fc-mediated effector functions, due to its high antigen-specific avidity and poor binding to Fcγ receptors (Marijn van der Neut Kolfschoten et al., 2007; Aurelia LC et al., 2025). To evaluate whether the emergence of IgG4 in vaccinated individuals impacts functional immunity, we assessed Fc-effector functions, specifically antibody-dependent cellular monocyte phagocytosis (ADCP), antibody-dependent neutrophil phagocytosis (ADNP), and antibody-dependent complement deposition (ADCD) across vaccine cohorts. Here we profiled the capacity of mRNA-1273-induced antibody responses (total IgG) to mediate antibody-dependent effector functions. Spike-specific antibodies response with ADCD activity was induced after the primary series, declined to near-baseline levels before the third dose, but were boosted to significantly higher levels post third immunization (log_2_ fold change baseline to post-prime = 2.38, and pre-boost to post-boost = 2.28) (**Figure 2A**). Spike-specific antibodies (total IgG) with ADCP activity were similarly robustly induced following the primary series, declined partially over time, and were boosted with the third dose, to a level comparable to post-prime responses (log_2_ fold change baseline to post-prime = 3.02, and pre-boost to post-boost = 0.72) (**Figure 2B**). Spike-specific antibodies (total IgG) with ADNP activity were significantly induced following the mRNA-1273 primary series, although with greater variability compared to the other functions. The ADNP activity declined over time and was subsequently boosted to higher levels after the third dose compared to the primary series (log_2_ fold change baseline to post-prime = 3.38, and pre-boost to post-boost = 2.39) (**Figure 2C**). Despite the significant increase in SARS-CoV-2-specific IgG4 antibody level, the functional properties of the vaccine-induced antibodies remain very robust. These results indicate that mRNA-1273 vaccination induce a multi-subclass antibody profile with robust Fc-effector functions at all timepoints evaluated.

**Figure 2.**
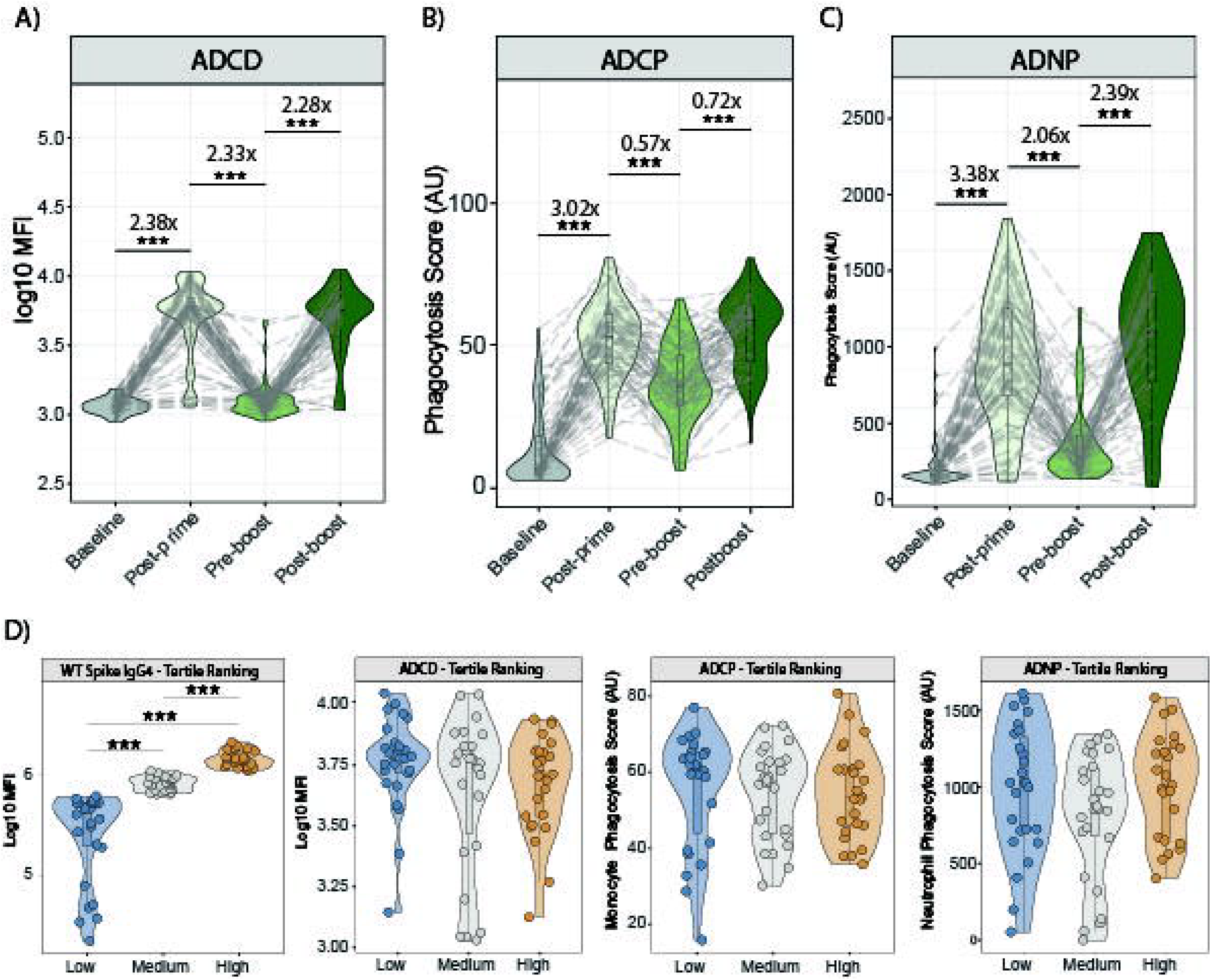
Functional Profiles of WT SARS-CoV-2 Spike-Specific IgG Across Vaccination Timepoints and IgG4 Tertiles. Violin plots show the ability of WT SARS-CoV-2 Spike-specific IgG antibodies to mediate antibody-dependent complement deposition (ADCD) **(A)**, antibody-dependent cellular phagocytosis (ADCP) **(B)**, and antibody-dependent neutrophil phagocytosis (ADNP) **(C)** across individuals (n = 79) vaccinated with mRNA-1273. Measurements were taken at baseline, post-prime (29 days after the second dose), pre-boost (11–14 months after the primary series), and post-boost (29 days after the booster). Dotted lines connect individual participants across all timepoints. Tertile analysis **(D)** groups participants based on their IgG4 antibody levels at the post-boost timepoint. The analysis divides the population into tertile 1 (lowest IgG4 antibody levels), tertile 2 (medium IgG4 antibody levels), and tertile 3 (highest IgG4 antibody levels) and compares ADCD, ADCP, and ADNP functionality across these groups. Mann–Whitney U-tests corrected for multiple comparisons using the Benjamini-Hochberg (BH) method were applied. Adjusted p-values are denoted as follows: ***p<0.001, **p<0.01, *p<0.05.

To rigorously examine the relationship between spike-specific IgG4 antibody levels and effector functions, we divided our cohort of vaccinated individuals into three tertiles based on low, medium, or high levels of spike-specific IgG4 antibodies post third immunization (**Figure 2D**). We then compared the levels of spike-specific total IgG antibodies displaying ADCD, ADCP, or ADNP activity across these three tertiles. The distribution of ADCD, ADCP, and ADNP activity appeared generally similar across the tertiles, with no evident skew or outliers within each group Overall, no statistically significant correlations were observed between the level of ADCD, ADCP and ADNP functions and the level of spike-specific IgG4 antibodies. These findings suggest that IgG4 antibody levels did not compromise Fc-effector functions in this cohort.

### mRNA-1273-induced IgG4 Antibody Responses Exhibited Enhanced Affinity and Neutralizing Activity

High antibody affinity is crucial for vaccine efficacy as it reflects the strength of binding between the antibodies and the virus, enhancing immune response durability and precision. Combined with potent neutralization, which directly blocks viral entry into host cells, high-affinity but non-neutralizing antibodies play a key role in mitigating disease severity in the context of SARS-CoV-2 (Clark JJ., 2024; Pierre CN. 2023; Zhang A., 2023). Previous studies have suggested that SARS-CoV-2 mRNA vaccines induce IgG responses with enhanced antibody affinity (Bellusci L., 2022). Therefore, to explore this further, we purified IgG4 antibodies from the serum of mRNA-1273 vaccinated individuals (n = 96) and measured spike-specific IgG4 antibody affinity before and after the third dose (booster). An affinity index was calculated by dividing the binding signal of IgG4 antibodies to spike with urea treatment by the binding signals of IgG4 antibodies to spike without urea treatment (**Figure 3A**). We observed a significant increase in the IgG4 affinity index following the third dose of mRNA-1273 compared to the pre-boost, confirming that purified IgG4 antibodies exhibited strong binding affinity to SARS-CoV-2 spike protein.

**Figure 3.**
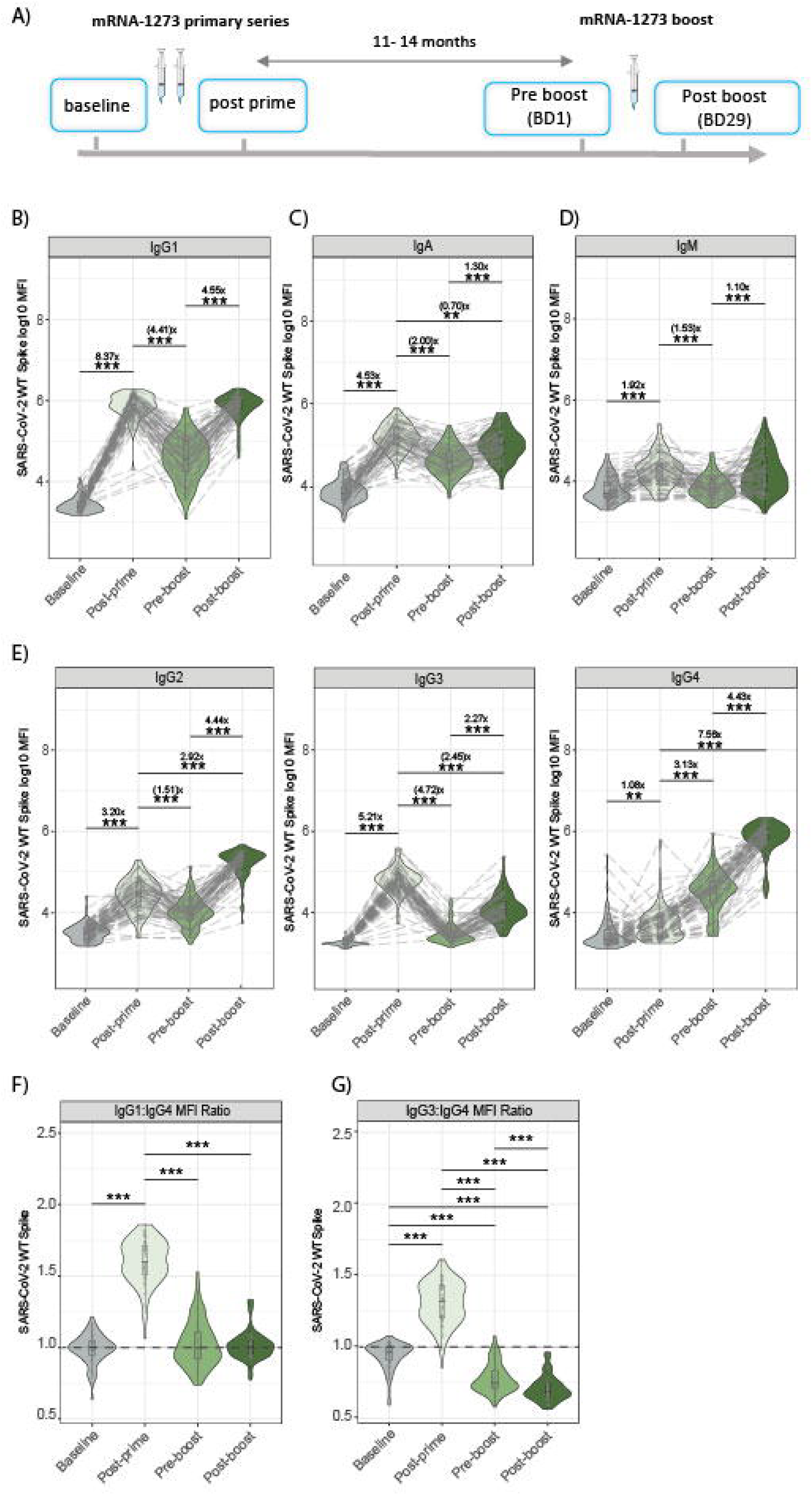
WT SARS-CoV-2 Spike-Specific IgG4 Affinity and Neutralization Capacity Pre- and Post-Boost. **(A)** Comparison of WT SARS-CoV-2 Spike-specific IgG4 antibody affinity pre- and post-boost. The relative affinity index was calculated as [IgG4 antibody level after urea treatment /IgG4 antibody level with PBS treatment] ×100. **(B)** Neutralization capacity of purified IgG4 antibodies normalized to 1 µg/ml of antibody (protein) measured by Nanodrop. **(C)** Neutralization capacity of total IgG antibodies normalized to 1 µg/ml of antibody. Dotted lines connect individual participants across all timepoints. Statistical significance was calculated using a Wilcoxon t-test, and fold changes were calculated for pre- and post-boost time points. P-values were adjusted using the Benjamini-Hochberg (BH) method and are denoted as follows:***p<0.001,**p<0.01, *p<0.05.

Neutralization assays are widely used as a surrogate for vaccine effectiveness and are considered one of the most clinically relevant measurement of protection. While it is well established that total IgG contributes to SARS-CoV-2 neutralization, the specific role of individual IgG subclasses, particularly IgG4, remains unclear. To address this gap, we assessed the neutralizing activity of purified IgG4 and total IgG antibodies from individuals vaccinated with mRNA-1273 using a SARS-CoV-2 pseudovirus neutralization assay (D614G). Serially diluted purified IgG4 and total IgG samples from individuals vaccinated with mRNA-1273 were incubated with SARS-CoV-2 pseudovirus (D614G), followed by infection of susceptible cells. Neutralization titers were determined as the reciprocal serum dilution required to achieve 50% inhibition of infection and normalized to 1 μg/mL of antibodies based on their concentration in each sample. Both purified IgG4 (**Figure 3B**) and total IgG antibodies (**Figure 3C**) showed a significant increase in neutralization capacity post-boost compared to pre-boost, in accordance with the observed increase in affinity correlated with enhanced neutralization. These findings suggest that high-affinity neutralizing IgG4 antibodies, resulting from continuous B-cell evolution, retain critical Fab-mediated functions that may contribute significantly to viral neutralization.

## Discussion

Antibodies serve as the primary correlate of protection for most approved vaccines (Plotkin, 2010; Plotkin, 2008). While IgG is the dominant antibody isotype in the blood, early IgM responses are commonly induced by viral or bacterial infections and various vaccine platforms. These IgM responses can variably class-switch to IgG (including IgG3, IgG1, IgG2, and IgG4) or IgA, depending on the vaccine platform, pathogen, or antigen type (Stavnezer J, 2008). Furthermore, the persistence of these isotypes following primary immunization and their behavior during boosting may vary between vaccine platforms (Serwanga J, 2024; Suwarti S, 2024). Protein alone, adjuvanted or vectored viral vaccines generally elicit IgG1/IgG3-biased humoral immune responses characterized by robust Fc-effector functions, while carbohydrate vaccines tend to induce IgM/IgG2-biased humoral responses (Barrett DJ, 1986; Stefanetti G, 2022). SARS-CoV-2 mRNA vaccines, however, present a unique immunological profile, characterized initially by a Th1-biased IgG1/IgG3 antibody response after the primary series (2 doses), which evolves into a multi-subclass and isotype response upon subsequent boosting (Irrgang, 2023; Gelderloos, 2024; Kiszel, 2023). This evolution is accompanied by enhanced Fcγ-mediated functions and distinct antibody dynamics that differ from traditional vaccine profiles.

The induction of IgG4 antibodies following repeated dosing of SARS-CoV-2 mRNA vaccination represents a novel and intriguing phenomenon. However, IgG4 class-switching is a natural response to repeated antigen exposure and has also been observed with other vaccine modalities (Neumeier, 2022; Hendrikx LH, 2011; Chaudhury S, 2017; Mdluli T, 2021; Kratochvil S, 2017). The IgG4 subclass is typically associated with immune tolerance and prolonged antigen exposure, differing substantially from other IgG subclasses in its functional properties (Aalberse, 2002). Unlike IgG1 and IgG3 antibodies, which are rapidly induced during primary immune responses and are highly pro-inflammatory, the delayed appearance of IgG4 antibodies likely reflects ongoing B-cell maturation and germinal center dynamics, emphasizing its role in long-term immune adaptation rather than immediate response. IgG4 antibodies are generally in low abundance (0–5% of total IgG) but may slowly increase over time due to repeated or excessive exposure to some antigens. Following repeated mRNA vaccination, IgG4 antibody level was observed to increase from 0.04% of total SARS-CoV-2 Spike–specific IgG after two doses (primary series) to 19.27% - 40% after third dose (boost) (Irrgang, 2023; Kiszel, 2023), with further increase post-fourth and fifth vaccination (Gelderloos, 2024). The IgG4 subclass is classically described as anti-inflammatory, with limited activation of Fc-mediated effector functions such as antibody-dependent cellular cytotoxicity, phagocytosis and complement activation (Rispens T. 2023).

While it has been speculated that increased IgG4 antibody levels may be associated with immunosuppression and poor clinical outcomes (Kalkeri R, 2024), this has not been observed with SARS-CoV-2 mRNA vaccines, which remain highly efficacious against symptomatic and severe COVID-19 (Figueroa AL, et al. 2025; Kopel H, et al. 2024; Baden L, et al. 2024; Berthaud V, et al. 2024; Figueroa A, et al. 2024; Sahly H. et al. 2021, Gilbert P. et al. 2022; Tseng, H.F, 2022;). This study shown that despite the significant rise in IgG4 antibody levels after mRNA-1273 vaccination, the overall functional antibody response remains robust, primarily due to highly Fc-functional IgG1 antibodies. Similarly, antibody-dependent cellular phagocytosis (ADCP) and antibody-dependent neutrophil phagocytosis (ADNP) responses are robustly enhanced. Additionally, IgG4 antibodies induced after a booster dose of mRNA-1273, exhibited high affinity for the SARS-CoV-2 Spike protein, maintaining effective neutralization capabilities. This high-affinity binding to the antigen ensures that IgG4 antibodies contribute to neutralization, reinforcing their functional importance despite reduced engagement with Fcγ receptors. The diminished interaction of IgG4 antibodies with Fcγ receptors and the inability to activate complement may reduce contribution to effector-mediated pathogen clearance. However, this property aligns with the broader role of the IgG4 subclass in minimizing inflammation during sustained antigen exposure. The balance between neutralization and reduced inflammation highlights the adaptability of IgG4 antibodies in fine-tuning the immune response to repeated vaccinations.

The late emergence of IgG4 antibodies after booster doses may reflect ongoing germinal center activity, favoring B-cell affinity maturation and a shift toward regulatory immune responses. Affinity maturation, where higher-affinity antibodies are selected over time through somatic hypermutation, enhances long-term immune protection (Chan and Brink, 2012). Continuous antigen presentation in germinal centers promotes this fine-tuning, underscoring the adaptability of the immune system to repeated antigen exposure. IgG4 antibodies may mitigate excessive inflammation, which could be beneficial in the context of repeated booster vaccinations. This phenomenon has potential implications for the design of booster strategies and next-generation vaccines aimed at optimizing both protective immunity and immune regulation. However, the kinetics and implications of maturing antibody affinity in the context of immune protection against SARS-CoV-2 and its variants of concern remain to be fully elucidated.

The emergence of IgG4 antibodies following repeated SARS-CoV-2 mRNA vaccination has raised concerns about potential decrease of vaccine-induced protection due to IgG4’s lower capacity for Fc-effector engagement. However, our results indicate that IgG4 antibodies, rather than suppressing immunity, retain or even potentiate functional antibody responses, likely due to affinity maturation and stable Fab-mediated viral neutralization. This suggests that contrary to the notion of IgG4 contributing to immune attenuation, IgG4 may actively participate in immune protection following SARS-CoV-2 mRNA vaccination. Importantly, this is consistent with real-world clinical observations showing that booster doses of SARS-CoV-2 mRNA vaccines continue to offer strong and incremental protection against COVID-19 (Tseng, H.F, 2022; Baden, L.R., 2024; Figueroa AL, et al. 2025).

In summary, the unique profile of IgG4 antibodies underscores the versatility of mRNA vaccines in shaping immune responses against SARS-CoV-2. The induction of IgG4 highlights the complexity of immune adaptation to repeated antigen exposure and suggests a broader functional role for IgG4 than previously appreciated. However, further research is necessary to elucidate its functional significance and to understand the long-term implications of IgG4-driven immunity for enhancing the efficacy and durability of mRNA-based vaccines against SARS-CoV-2 and other pathogens.

## Data Availability

Access to patient-level data presented in this article and supporting clinical documents with external researchers who provide methodologically sound scientific proposals will be available upon reasonable request for products or indications that have been approved by regulators in the relevant markets and subject to review from 24 months after study completion. Such requests can be made to Moderna Inc., 325 Binney Street, Cambridge, MA, 02142 USA ≤ ≥. A materials transfer and/or data access agreement with the sponsor will be required for accessing shared data. All other relevant data are presented in the paper.

## Notes

### Competing Interest Statement

PK, HB, DL, DC, W-HY, KW, SC, RW, DE, and CH are employees of Moderna, Inc., and hold stock/stock options in the company. GA is a former employee of Moderna Inc.

### Funding Statement

This study was funded by Moderna, Inc.

### Author Declarations

Advarra, Columbia, MD, USA

## References

Aalberse RC, Schuurman J. IgG4 breaking the rules. Immunology. 2002 Jan;105(1):9–19. doi: 10.1046/j.0019-2805.2001.01341.x. PMID: 11849310; PMCID: PMC1782638.

Ackerman ME, Moldt B, Wyatt RT, Dugast AS, McAndrew E, Tsoukas S, Jost S, Berger CT, Sciaranghella G, Liu Q, Irvine DJ, Burton DR, Alter G, A robust, high-throughput assay to determine the phagocytic activity of clinical antibody samples, J. Immunol. Methods (2011), doi: 10.1016/j.jim.2010.12.016.

Aurelia LC et al,. Increased SARS-CoV-2 IgG4 has variable consequences dependent upon Fc function, Fc receptor polymorphism, and viral variant. Sci Adv. 2025 Feb 28;11(9):eads1482. doi: 10.1126/sciadv.ads1482. Epub 2025 Feb 26. PMID: 40009690; PMCID: PMC11864192.

Baden, L.R., El Sahly, H.M., Essink, B. et al. Long-term safety and effectiveness of mRNA-1273 vaccine in adults: COVE trial open-label and booster phases. Nat Commun 15, 7469 (2024). 10.1038/s41467-024-50376-z

Berthaud V, et al. Safety and Immunogenicity of an mRNA-1273 Booster in Children. Clin Infect Dis. 2024 Dec 17;79(6):1524–1532. doi: 10.1093/cid/ciae420. PMID: 39158584; PMCID: PMC11650855.

Barrett DJ, Ayoub EM. IgG2 subclass restriction of antibody to pneumococcal polysaccharides. Clin Exp Immunol. 1986 Jan;63(1):127-34. PMID: 3955880; PMCID: PMC1577346.

Bates TA, et al. BNT162b2-induced neutralizing and non-neutralizing antibody functions against SARS-CoV-2 diminish with age. Cell Rep. 2022 Oct 25;41(4):111544. doi: 10.1016/j.celrep.2022.111544. Epub 2022 Oct 5. PMID: 36252569; PMCID: PMC9533669.

Bellusci, L., Grubbs, G., Zahra, F.T. et al. Antibody affinity and cross-variant neutralization of SARS-CoV-2 Omicron BA.1, BA.2 and BA.3 following third mRNA vaccination. Nat Commun 13, 4617 (2022). 10.1038/s41467-022-32298-w

Boudreau CM, Yu WH, Suscovich TJ, Talbot HK, Edwards KM, Alter G, Selective induction of antibody effector functional responses using MF59-adjuvanted vaccination, J. Clin. Invest. (2020), doi: 10.1172/JCI129520.

Brown EP, Dowell KG, Boesch AW, Normandin E, Mahan AE, Chu T, Barouch DH, Bailey-Kellogg C, Alter G, Ackerman ME, Multiplexed Fc array for evaluation of antigen-specific antibody effector profiles, J. Immunol. Methods (2017), doi: 10.1016/j.jim.2017.01.010.

Chalkias S, et al. Safety and Immunogenicity of a 100 μg mRNA-1273 Vaccine Booster for Severe Acute Respiratory Syndrome Coronavirus-2 (SARS-CoV-2). medRxiv 2022;:2022.03.04. 22271830. doi: 10.1101/2022.03.04.22271830. PMID: 35291289; PMCID: PMC8923111.

Chan TD, Brink R. Affinity-based selection and the germinal center response. Immunol Rev. 2012;247(1):11–23. doi: 10.1111/j.1600-065X.2012.01118.x

Chen X, et al., Neutralizing Antibodies Against Severe Acute Respiratory Syndrome Coronavirus 2 (SARS-CoV-2) Variants Induced by Natural Infection or Vaccination: A Systematic Review and Pooled Analysis, Clinical Infectious Diseases, Volume 74, Issue 4, 15 February 2022, Pages 734–742, doi.org/10.1093/cid/ciab646

Chaudhury S, et al. Delayed fractional dose regimen of the RTS,S/AS01 malaria vaccine candidate enhances an IgG4 response that inhibits serum opsonophagocytosis. Sci Rep. 2017 Aug 11;7(1):7998. doi: 10.1038/s41598-017-08526-5. PMID: 28801554; PMCID: PMC5554171.

Clark JJ, et al. Protective effect and molecular mechanisms of human non-neutralizing cross-reactive spike antibodies elicited by SARS-CoV-2 mRNA vaccination. Cell Rep. 2024 Nov 26;43(11):114922. doi: 10.1016/j.celrep.2024.114922. Epub 2024 Nov 5. PMID: 39504245; PMCID: PMC11804229.

Davis CW et al. Longitudinal Analysis of the Human B Cell Response to Ebola Virus Infection. Cell. 2019, 177(6):1566-1582.e17. doi: 10.1016/j.cell.2019.04.036. PMID: 31104840; PMCID: PMC6908968.

Figueroa AL, et al. Safety and immunogenicity of an mRNA-1273 vaccine booster in adolescents. Hum Vaccin Immunother. 2025 Dec;21(1):2436714. doi: 10.1080/21645515.2024.2436714

Figueroa AL, et al. Safety and durability of mRNA-1273-induced SARS-CoV-2 immune responses in adolescents: results from the phase 2/3 TeenCOVE trial. EClinicalMedicine. 2024 Jul 18;74:102720. doi: 10.1016/j.eclinm.2024.102720. PMID: 39091673; PMCID: PMC11293523.

Follmann D, et al. Who to Boost When: The Effect of Age and Dosing Interval on Delta and Omicron COVID-19 Incidence in the Open-label Phase of the COVE Trial. Open Forum Infect Dis. 2024 Nov 25;11(12):ofae689. doi: 10.1093/ofid/ofae689. PMID: 39679349; PMCID: PMC11639572.

Gelderloos, A.T., Verheul, M.K., Middelhof, I. et al. Repeated COVID-19 mRNA vaccination results in IgG4 class switching and decreased NK cell activation by S1-specific antibodies in older adults. Immun Ageing 21, 63 (2024). 10.1186/s12979-024-00466-9

Gilbert PB, et al. Immune correlates analysis of the mRNA-1273 COVID-19 vaccine efficacy clinical trial. Science. 2022 Jan 7;375(6576):43–50. doi: 10.1126/science.abm3425. Epub 2021 Nov 23. PMID: 34812653; PMCID: PMC9017870.

Hendrikx LH, et al. Different IgG-subclass distributions after whole-cell and acellular pertussis infant primary vaccinations in healthy and pertussis infected children. Vaccine. 2011 Sep 16;29(40):6874–80. doi: 10.1016/j.vaccine.2011.07.055. Epub 2011 Jul 29. PMID: 21803088.

Irrgang P; et al. Class switch toward noninflammatory, spike-specific IgG4 antibodies after repeated SARS-CoV-2 mRNA vaccination. Sci Immunol. 2023 Jan 27;8(79):eade2798. doi: 10.1126/sciimmunol.ade2798. Epub 2023 Jan 27. PMID: 36548397; PMCID: PMC9847566.

Kalkeri R.; et al. Altered IgG4 antibody response to repeated mRNA versus recombinant protein SARS-CoV-2 vaccines. 2024, Journal of Infection, Volume 88, Issue 3, 106119

Kaplonek P. et al., mRNA-1273 vaccine-induced antibodies maintain Fc effector functions across SARS-CoV-2 variants of concern, Immunity 2022, Volume 55, Issue 2, ISSN 1074-7613, 10.1016/j.immuni.2022.01.001

Karsten CB, Mehta N, Shin SA, Diefenbach TJ, Slein MD, Karpinski W, Irvine EB, Broge T, Suscovich TJ, Alter G, A versatile high-throughput assay to characterize antibody-mediated neutrophil phagocytosis, J. Immunol. Methods (2019), doi: 10.1016/j.jim.2019.05.006.

Kiszel P., Sík, P., Miklós, J. et al. Class switch towards spike protein-specific IgG4 antibodies after SARS-CoV-2 mRNA vaccination depends on prior infection history. Sci Rep 13, 13166 (2023). 10.1038/s41598-023-40103-x

Kopel H, et al. Effectiveness of the 2023-2024 Omicron XBB.1.5-containing mRNA COVID-19 Vaccine (mRNA-1273.815) in Preventing COVID-19-related Hospitalizations and Medical Encounters Among Adults in the United States. Open Forum Infect Dis. 2024 Nov 26;11(12):ofae695. doi: 10.1093/ofid/ofae695. PMID: 39691287; PMCID: PMC11651145.

Kratochvil S, et al. Phase 1 Human Immunodeficiency Virus Vaccine Trial for Cross-Profiling the Kinetics of Serum and Mucosal Antibody Responses to CN54gp140 Modulated by Two Homologous Prime-Boost Vaccine Regimens. Front Immunol. 2017 May 24;8:595. doi: 10.3389/fimmu.2017.00595. PMID: 28596770; PMCID: PMC5442169.

Lin DY, et al. Effectiveness of Bivalent Boosters against Severe Omicron Infection. N Engl J Med. 2023 Feb 23;388(8):764–766. doi: 10.1056/NEJMc2215471. Epub 2023 Jan 25. PMID: 36734847; PMCID: PMC9933929.

Marijn van der Neut Kolfschoten et al., Anti-Inflammatory Activity of Human IgG4 Antibodies by Dynamic Fab Arm Exchange. Science 317,1554–1557 (2007). DOI:10.1126/science.1144603

Mdluli T, et al. RV144 HIV-1 vaccination impacts post-infection antibody responses. PLoS Pathog. 2021 Mar 2;17(3):e1009386. doi: 10.1371/journal.ppat.1009386. PMID: 33651828; PMCID: PMC7924741.

Mistry P et al, (2022) SARS-CoV-2 Variants, Vaccines, and Host Immunity. Front. Immunol. 12:809244. doi: 10.3389/fimmu.2021.809244

Moderna Press Release 2020 https://investors.modernatx.com/news/news-details/2020/Moderna-Announces-Primary-Efficacy-Analysis-in-Phase-3-COVE-Study-for-Its-COVID-19-Vaccine-Candidate-and-Filing-Today-with-U.S.-FDA-for-Emergency-Use-Authorization/default.aspx

Neumeier D, et al. Phenotypic determinism and stochasticity in antibody repertoires of clonally expanded plasma cells. Proc Natl Acad Sci U S A. 2022 May 3;119(18):e2113766119. doi: 10.1073/pnas.2113766119. Epub 2022 Apr 29. PMID: 35486691; PMCID: PMC9170022.

Pfizer Press Release 2020,https://www.pfizer.com/news/press-release/press-release-detail/pfizer-and-biontech-conclude-phase-3-study-covid-19-vaccine.

Pierre CN, et al. Non-neutralizing SARS-CoV-2 N-terminal domain antibodies protect mice against severe disease using Fc-mediated effector functions. PLoS Pathog. 2024 Jun 20;20(6):e1011569. doi: 10.1371/journal.ppat.1011569. PMID: 37546738; PMCID: PMC10402036.

Plotkin, SA. Correlates of protection induced by vaccination. Clinical and Vaccine Immunology. 2010, 17(7), 1055–1065. 10.1128/CVI.00021-10

Plotkin SA. Correlates of vaccine-induced immunity. Clinical Infectious Diseases. 2008;47(3):401-409. doi:10.1086/589862.

Pollard, A.J., Bijker, E.M. A guide to vaccinology: from basic principles to new Developments. Nat Rev Immunol 21, 83–100 (2021). 10.1038/s41577-020-00479-7

Rispens T, Huijbers MG. The unique properties of IgG4 and its roles in health and disease. Nat Rev Immunol. 2023 Nov;23(11):763–778. doi: 10.1038/s41577-023-00871-z. Epub 2023 Apr 24. PMID: 37095254; PMCID: PMC10123589.

El Sahly HM, et al. COVE Study Group. Efficacy of the mRNA-1273 SARS-CoV-2 Vaccine at Completion of Blinded Phase. N Engl J Med. 2021 Nov 4;385(19):1774–1785. doi: 10.1056/NEJMoa2113017. Epub 2021 Sep 22. PMID: 34551225; PMCID: PMC8482810.

Serwanga J, et al. Persistent and robust antibody responses to ChAdOx1-S Oxford-AstraZeneca (ChAdOx1-S, Covishield) SARS-CoV-2 vaccine observed in Ugandans across varied baseline immune profiles. PLoS ONE 2024; 19(7): e0303113

Stavnezer J, Guikema JE, Schrader CE. Mechanism and regulation of class switch recombination. Annu Rev Immunol. 2008;26:261-92.doi: 10.1146/annurev.immunol.26.021607.090248. PMID: 18370922; PMCID: PMC2707252.

Stefanetti G, et al. Immunobiology of Carbohydrates: Implications for Novel Vaccine and Adjuvant Design Against Infectious Diseases. Front Cell Infect Microbiol. 2022 Jan 18;11:808005. doi: 10.3389/fcimb.2021.808005. PMID: 35118012; PMCID: PMC8803737

Suwarti, S., Lazarus, G., Zanjabila, S. et al. Anti-SARS-CoV-2 antibody dynamics after primary vaccination with two-dose inactivated whole-virus vaccine, heterologous mRNA-1273 vaccine booster, and Omicron breakthrough infection in Indonesian health care workers. BMC Infect Dis 2024 24, 768. 10.1186/s12879-024-09644-y

Tseng, H.F., Ackerson, B.K., Luo, Y. et al. Effectiveness of mRNA-1273 against SARS-CoV-2 Omicron and Delta variants. Nat Med 2022, 28, 1063–1071. 10.1038/s41591-022-01753-y

Zhang A, et al. Beyond neutralization: Fc-dependent antibody effector functions in SARS-CoV-2 infection. Nat Rev Immunol. 2023 Jun;23(6):381–396. doi: 10.1038/s41577-022-00813-1. Epub 2022 Dec 19. PMID: 36536068; PMCID: PMC9761659.

Zhang, B., Fong, Y., Fintzi, J. et al. Omicron COVID-19 immune correlates analysis of a third dose of mRNA-1273 in the COVE trial. Nat Commun 15, 7954 (2024). 10.1038/s41467-024-52348-9

